# Profiling abuse and neglect of women with disabilities: a step towards prevention of mistreatment of vulnerable populations

**DOI:** 10.1101/2023.07.27.23293278

**Authors:** Josephine Savard, Georgios Gavriilidis, Anna Lindblad, Jesse Huang, Milena Zeitelhofer Adzemovic

**Affiliations:** Department of Clinical Neuroscience, Center for Molecular Medicine, Karolinska Institutet, Stockholm, Sweden; Stockholm Centre for Healthcare Ethics, Department of Learning, Informatics, Management and Ethics, Karolinska Institutet, Stockholm, Sweden

## Abstract

Women with disabilities are at increased risk of violence and neglect, and the physical and psychological barriers to seeking help often lead to prolonged periods of abuse. In addition to being a leading cause of acute injuries and numerous chronic diseases, exposure to violence also negatively affects mental health.

In this cross-sectional quantitative data analysis on experiences of violence among women with physical disabilities resulting from cerebral palsy (CP), multiple sclerosis (MS), traumatic brain injury (TBI), stroke and arthritis, a high prevalence of abuse and neglect can be confirmed. These groups were also compared with women who have visual- and hearing impairment. We could observe that type of mistreatment, perpetrators and required personal assistance differ between disability groups. Interestingly, the highest frequency of abuse was observed among women with hearing impairment, including number of ongoing incidences at the time of response. Moreover, denial of help with basic needs or prevented use of assistive devices was again more commonly associated with hearing impairment but also with MS. Since it has been shown that hearing impairment is related to the risk for cognitive decline and one of the greatest risks for dementia, it is tempting to speculate that cognitive impairment may not only enhance but pose a higher risk factor for abuse than physical disability itself, hence calling for further research and more targeted interventions to prevent violence and support victims among women with disabilities.

## Introduction

Women with disabilities have a higher risk of being exposed to neglect and violence, which often differ by the duration of abuse and relationship with the perpetrator^1,2^. Exposure to violence may also be associated with other risk factors, such as low socioeconomic status and education, which are not only related to unemployment and economic dependence but isolation and substance abuse^3-6^. Violence is also associated with many perpetrator-related characteristics, such as excessive alcohol consumption and patriarchal dominance, particularly jealousy and possessive behavior^3,7^. Of note, violence is consistently a leading cause of acute injury among women^4^ and often correlates with worse long-term health outcomes, including increased risk of many chronic and stress-related diseases^8-10^. It also negatively impacts mental health with higher rates of psychiatric disorders, including depression, generalized anxiety disorder, post-traumatic stress disorder, and drug and alcohol dependence^6,10^. In addition, disabilities pose physical and psychological barriers to seeking help, often leading to extended periods of abuse^10,12^.

Although societal concerns about abuse among women with disabilities have increased, the extent of neglect and violence has not been adequately investigated. This is particularly applying to the lack of evidence regarding types of mistreatments in different disability/diagnostic groups. Building on the previous study by Milberger et al^12^, we cross-examine the characteristics of abuse associated with physical disabilities of different medical conditions, including cerebral palsy (CP), multiple sclerosis (MS), traumatic brain injury (TBI), stroke, and arthritis, in addition to general hearing and visual impairments. Our findings provide further insight into the types of abuse related to certain disability groups, which can help develop more targeted strategies to address mistreatment in vulnerable populations.

## Method

The initial study by Milberger et al. was approved by the Investigative Review Board at Wayne State University, and the dataset used is publicly available; the study details and protocols have been described previously^12^. In summary, women (≥18 years old) with physical disabilities were recruited through various organizations primarily servicing those with physical disabilities, across the state of Michigan (USA) between 2000 and 2001. An initial interview was conducted either by phone or a self-administered questionnaire concerning any history of physical or sexual abuse, along with incidences where an individual prevented the use of an assistive device (e.g., wheelchair, cane, respirator) or refused to help with basic personal needs (e.g., medication, personal hygiene). Of the 177 enrolled participants, 100 women (56%) reported experiencing abuse, of which 85 women responded to a follow-up questionnaire detailing the history and relationships associated with their abuse.

The present study extends towards evaluation of types of abuse among women with disabilities resulting from CP (n= 33), MS (n= 22), arthritis (n= 43), TBI (n= 13), and stroke (n= 8). These groups were also compared against women with visual (n= 28) and hearing impairment (n= 29). Individuals with other disabilities, such as spina bifida, lupus, and post-polio, or not of Caucasian or African American ethnicities, were excluded from this analysis due to the limited sample size. In summary, 130 participants at baseline were included in our analysis, with their demographic characteristics summarized in **Table 1**. Of note, some of the study participants were eligible for more than one diagnostic/disability group. Differences in the frequency of measure for abuse and physical impairment between groups were assessed using Fischer’s exact test. All analyses and illustrations were performed with R(v.4.2.1).

**Table 1.**
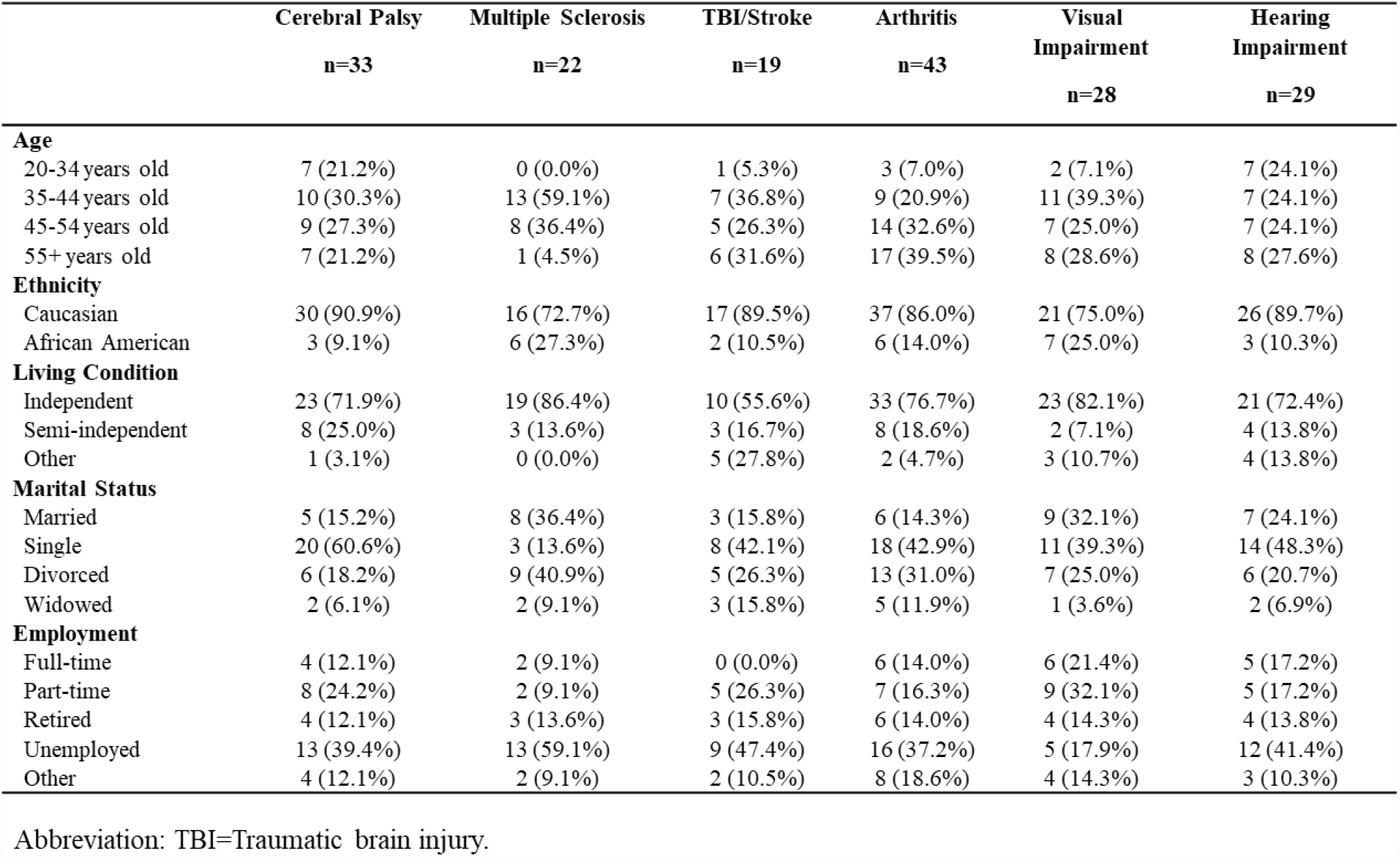
Descriptive statistics of cohort, stratified by disability type.

## Results

Study participants were primarily between the ages of 35-54 at the time of enrollment, except for those with arthritis who were generally older (45+ years old). Most women were unemployed or retired (53.8%), with the highest unemployment rate among women with MS (59.1%). Many also lived independently (78.1%), single or divorced at enrollment, with more than half requiring personal assistance (56.9%, **Figure 1a**). In particular, women with CP were more likely to require assistance services for dressing (30.3%, p=0.01), toileting (18.2%, p=0.02), personal hygiene (30.3%, p=0.008), and meal preparation (42.4%, p=0.01). Assistance with home maintenance was more common among women with TBI and stroke (78.9%, p=0.0009), while those with hearing impairment were more likely to require assistance with taking medication (27.6%, p=0.04).

**Figure 1.**
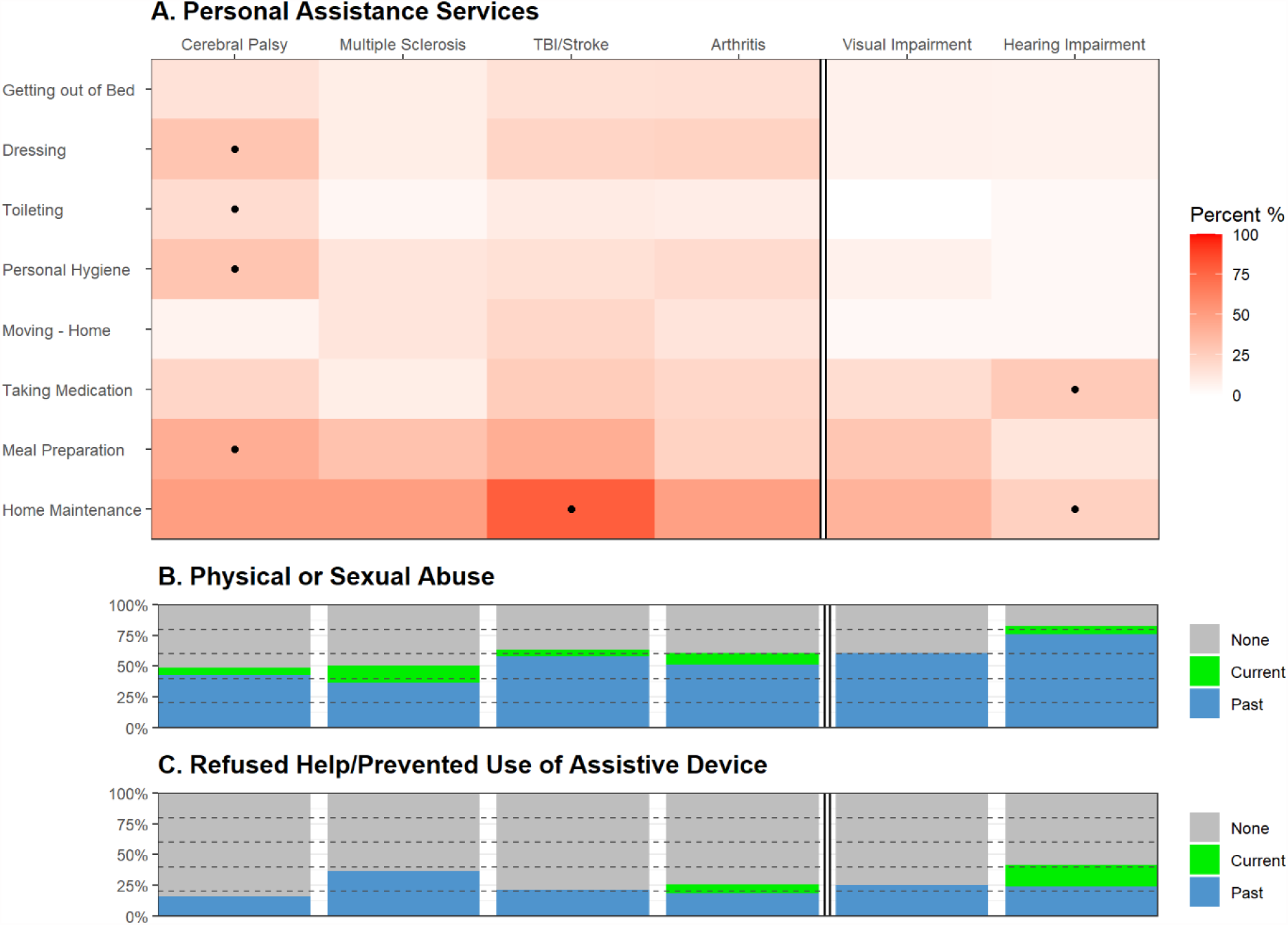
Physical and sexual abuse among women with disabilities. Heatmap (A) illustrates frequency rate (%) of self-reported use of personal assistance services. Frequencies were compared between disability groups using Fisher’s exact test with P<0.05 highlighted (dot). Stacked barplots illustrate the frequency rate (%) of [B] physical or sexual abuse and [C] incidences when the depended individual refused to help with a personal need or prevented the use of an assistive device (current, past, or none). Abbreviation: TBI=Traumatic Brain Injury

Most women (57.7%) have experienced physical or sexual abuse as adults, with 7.7% reporting ongoing abuse at the time of response (**Figure 1b**). Interestingly, the highest frequency of abuse was observed among those with hearing impairment (82.8%, p=0.002), followed by TBI and stroke (63.2%), visual impairment (60.7%), arthritis (60.5%), MS (50.0%), and lastly CP (48.5%). Approximately a quarter of the study participants also reported being denied help with basic needs or prevented from using assistive devices (**Figure 1c**). This was more commonly associated with hearing impairment (41.4%, p=0.03) and MS (36.3%). Notably, more women with hearing impairment reported ongoing incidences at the time of response (17.2%, p=0.006). Although the causes of visual and hearing impairment were unspecified, women with hearing impairments had a slightly higher proportion of arthritis, indicating age may play a role in some cases. Similarly, women with visual impairments had a greater frequency of neurological conditions, including MS (13.3%), TBI (13.3%), and spinal cord injury (6.7%). However, these trends were merely suggestive of the baseline characteristics of the study participants.

Among those that responded to the follow-up questionnaire regarding their history of abuse (n= 57), most women reported physical/sexual abuse either from a partner (89.6%/62.9%) or family members (35.4%/17.1%) rather than strangers (4.2%/11.4%). As shown in **Figure 2**, women with CP had a higher risk of physical abuse and being denied assistance with basic needs from healthcare providers (p<0.05). This is likely related to the frequent need for more essential personal assistance (e.g., dressing, toileting, and personal hygiene) compared with other disabilities (**Figure 1**). On the contrary, women with MS, arthritis, and visual impairment, were more likely to experience abuse by a partner (p=0.02).

**Figure 2.**
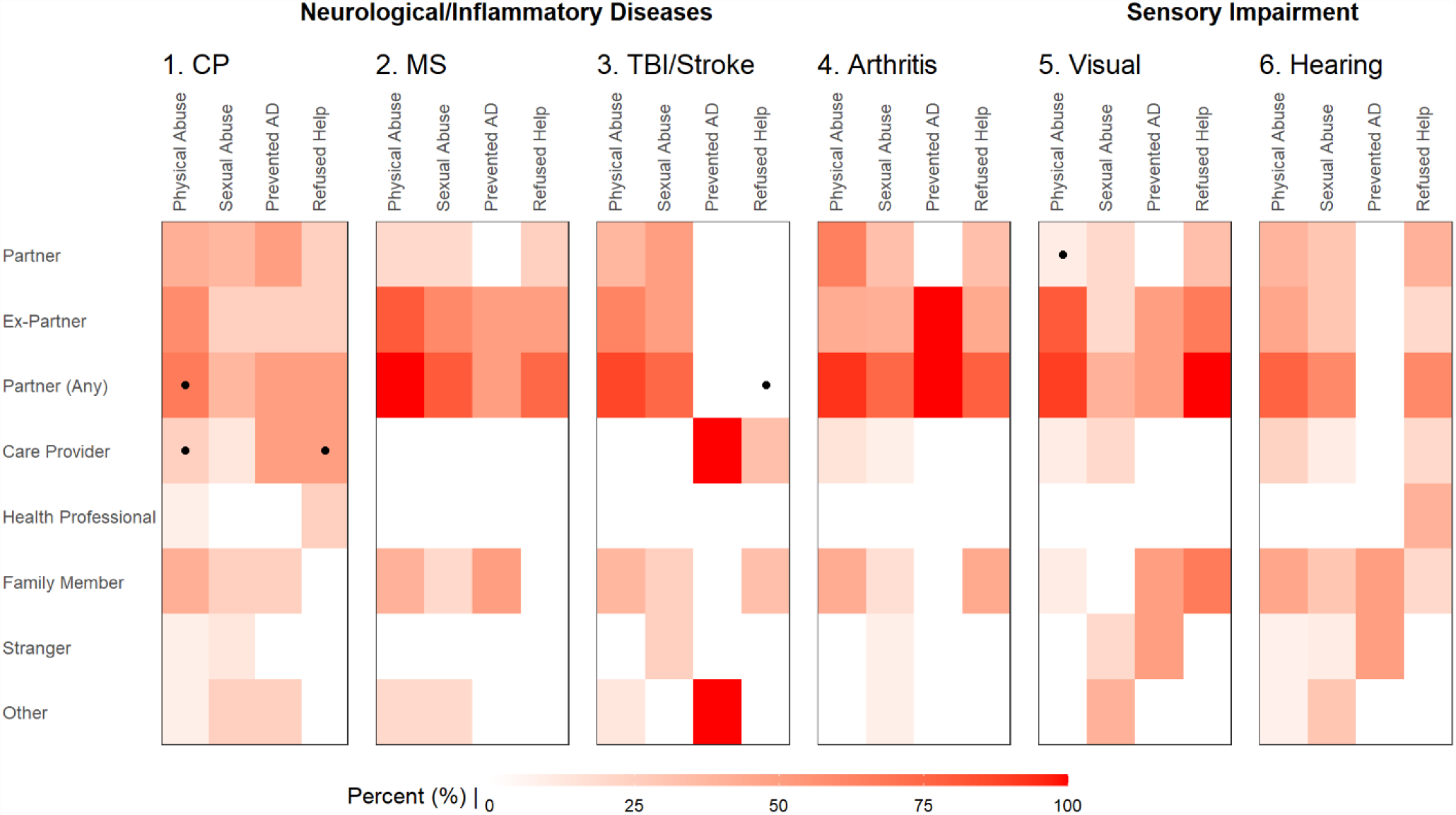
Comparing the frequency of relationships with perpetrators among women with different disabilities. Heatmap illustrating the reported frequency rate (%) of perpetrators among those experiencing physical abuse, sexual abuse, prevented use of assistive device (AD), and refused help of basic needs. Frequencies were compared between diseases groups using Fisher’s exact test with P<0.05 highlighted (dot). Abbreviations: CP=Cerebral palsy, MS=multiple sclerosis, TBI=traumatic brain injury.

## Discussion

Our findings indicate that nature of disability shapes the type of abuse, which is consistent with previous studies^13^. These differences are likely dependent on the type and severity of functional impairment and its required assistive services^10,14^. Progressive diseases, such as MS and rheumatic conditions, may also affect the risk and duration of abuse through psychological and social factors relating to the management of disease activity^10^. Furthermore, the timing of disability onset and its progression can affect an individual’s reliance on and susceptibility to abuse from certain perpetrators (e.g., care providers, intimate partners, and family members)^14^. These factors may also lead to sampling bias as women with severe or acute physical disabilities are often underrepresented^16,18^.

In this study, we could observe that women with CP were more often physically abused and refused assistance from care providers, likely due to their frequent interactions with the personnel, and a higher degree of personal care required^14^. However, this was not the case for less debilitating conditions, where violence and neglect were predominantly perpetrated by intimate partners^1,4,10,11^.

Interestingly, women with hearing impairment did not differ in type of abuse relative to other conditions. Nevertheless, they had the highest overall frequency of abuse, including more ongoing incidents at the time of response. Since deafness is frequently associated with muteness and impaired articulation, those with hearing impairment may be more susceptible to violence and remain in abusive relationships due to difficulties communicating and seeking help^15^. Notably, women with hearing problems were generally less likely to need personal assistance, which is commonly associated with a higher risk for interpersonal violence. Instead, they were more likely to require assistance with their medication. This may be attributed to an altered perception of time or oblivion associated with the reduction of external stimuli, but also to a cognitive decline. In this context, it has been shown that hearing impairment is related to the risk for cognitive decline, brain atrophy and tau accumulation^21^, and considered one of the greatest risks for dementia^22^. Hence, based on our findings, it is tempting to speculate that cognitive impairment may not only enhance, but pose a higher risk factor for abuse than physical disability itself, thus calling for further research.

This study evaluates the relationship of abuse among Caucasian and African-American women living in the USA. However, the associative profile among women of other countries and ethnic groups requires further investigation, as cultural barriers and sociodemographic differences influence the perception of abuse and disabilities^3,16,17^. Several confounding risk factors include living arrangements, education, poverty, and perpetuator-related characteristics, particularly substance abuse^4,7^. However, due to limitations in study power, we could not assess the detailed interactions between subtypes and severities of abuse with other demographic and lifestyle factors.

In conclusion, the present study confirms the high prevalence of abuse and neglect among women with disabilities, calling for further research to address patterns of violence specific to certain disability subtypes^10,18,19^. Such knowledge could facilitate more targeted interventions to support this heterogeneous population^13,18^. However, many victims report barriers to seeking help, which is pronounced in women with disabilities due to functional limitations and the emphasized perception of shame, guilt, and inferiority^10,12,17^. Screening strategies customized to common high-risk disability groups may improve the detection of ongoing victimization and ease access to support^1,8,11,19,20^. Lastly, further investigations must identify the long-term physical, social, and psychological implications of abuse^6,8^ and how that relates to standard clinical care of disabilities and their underlying medical conditions.

## Data Availability

All data produced in the present study are available upon reasonable request to the authors.

## Data availability

Data and related documentation from this study are stored and accessible through the Michigan government public repository. Additional summary data and analysis scripts can be made available by request.

## Acknowledgments

The authors thank the participants and professionals contributing to the data collection. The authors especially thank Sharon Milberger, ScD, for generously providing the original raw dataset.

